# Effects of an online plain language tool on health information quality: A randomised controlled trial

**DOI:** 10.1101/2024.04.04.24305365

**Authors:** Julie Ayre, Carissa Bonner, Danielle M Muscat, Erin Cvejic, Olivia Mac, Dana Mouwad, Heather L Shepherd, Parisa Aslani, Adam G Dunn, Kirsten J McCaffery

## Abstract

**Objective:** Complex and ineffective health communication is a critical and persistent source of inequity in our health systems. This occurs despite repeated policy directives to provide patients and community with health information that is easy to understand and that applies health literacy principles. This study sought to evaluate the effectiveness of the Sydney Health Literacy Lab (SHeLL) Health Literacy Editor, an easy-to-use online plain language tool that supports health information providers to apply health literacy guidelines to written health information.

**Design:** Randomised controlled trial with analysts blind to intervention group.

**Setting:** Online study, Australia

**Participants:** 188 health information providers with no previous experience using the Health Literacy Editor (mean age 41.0 (SD=11.6); 154 female (85%)).

**Intervention:** Participants were provided access to the Health Literacy Editor and a 30-minute online training program prior to editing three pre-specified health texts. The Health Literacy Editor gives objective, real-time, and fine-grained feedback on words and sentences. Control participants were asked to revise the texts using their own standard health information development processes.

**Main outcome measure:** Pre-registered primary outcome was text grade reading score (using validated instrument, the Simple Measure of Gobbledygook). Secondary outcomes were use of complex language (% of the text) and passive voice (number of instances), subjective expert ratings (Patient Education Materials Assessment Tool), and acceptability ratings (System Usability Scale; Technology Acceptance Model).

**Results:** Texts revised in the intervention group had significantly improved grade reading scores relative to control (Mean Difference (MD)=2.48, 95% CI=1.84 to 3.12, p<0.001, d=0.99), lower text complexity (MD=6.86, 95% CI=4.99 to 8.74, p<0.001, d=0.95) and less passive voice (MD=0.95, 95% CI=0.4 to 1.5, p<0.001, d=0.53) in intention-to-treat analyses. Experts rated texts in the intervention group more favourably for word choice and style than those in the control group (MD=0.44, 95% CI=0.25 to 0.63, p<0.001, d=0.63), with no loss of meaning or content. Participants rated the Health Literacy Editor an acceptable product (71.0/100, SD=13.7) that was useful (3.8/5, SD=0.7) and easy to use (4.0/5, SD=0.6).

**Conclusions and relevance:** The Health Literacy Editor helped users simplify health information and apply health literacy guidelines to written text. It has high potential to improve development of health information for people who have low health literacy. As an online tool the Health Literacy Editor is also easy to access and implement at scale.

**Trial registration:** ACTRN12623000386639

**Summary box:** *Section 1: What is already known on this topic?:*

- Most health information is hard for people to understand, particularly those who are older, with less education, or who speak English as a second language.
- Systematic reviews show that texts that follow health literacy guidelines (e.g. use simpler words, shorter sentences and active voice) are easier for people to understand and recall.
- There are few automated tools that guide development of easy-to-understand written health information and none that have been rigorously evaluated in a randomised controlled trial.

*Section 2: What this study adds:*

- Participants who used the Health Literacy Editor were able to more effectively simplify health information compared to participants in the control group.
- On average participants in the intervention group produced texts suitable for a person with almost 2.5 fewer years of school education compared to those in the control group. Similar patterns were observed for complex language and passive voice.
- The Health Literacy Editor is an effective tool to support development of written health information that adheres to plain language principles. It can be used in clinical and non-clinical settings and implemented at scale.

---

National and international policies recognise that health literacy, a person’s capacity to access, understand, and act on health information, is a critical source of inequity in our health systems (1–3). Low health literacy contributes to higher mortality, morbidity, rates of hospitalisation, emergency department visits, and medication errors, independently of other social determinants such as age, education, and socioeconomic disadvantage (4). A key feature of these policies is the directive to provide health information that all people can easily understand, including people with low health literacy (5). There has been a failure to systematically integrate such directives into routine public health and clinical practice, despite some of these policies existing for over a decade (6–11).

To support the provision of easy-to-understand health information, there are several freely available and comprehensive guidelines that provide advice about how to apply health literacy and plain language principles to health information (12–16). These guidelines recommend evidence-based strategies to improve knowledge and recall of health information, such as putting essential information first, using simple language, and minimising medical jargon (17, 18). However, accompanying systems, training, and tools are needed to drive meaningful change in health literacy practices within an organisation (19, 20).

Online tools are well placed to help improve the application of health literacy guidelines because of their capacity to provide specific, immediate and actionable feedback on written text (21–23). These tools typically identify difficult words, sentences, and grammatical structures, and sometimes integrate technologies such as natural language processing and artificial intelligence (24–30). However, few have been specifically designed for health contexts (28–30), only two have been formally evaluated, and both evaluations were limited by small sample size and pre-post study design (29, 30). Though these tools hold promise, it is unclear how effectively health information providers incorporate the tool’s feedback when revising and simplifying health information.

To address this gap in the research, the current study aimed to evaluate whether using the ‘Sydney Health Literacy Lab (SHeLL) Health Literacy Editor’ (referred to herein as ‘Health Literacy Editor’) can support health information providers to effectively apply health literacy guidelines to written health information. The Health Literacy Editor is a new online tool that provides objective and immediate assessment of written health information across a range of factors, including feedback on school grade reading levels, complex language, passive voice, text structure, lexical density and diversity, and person-centred language (31). The Health Literacy Editor guides users in real-time, providing simpler or more familiar alternatives for medical and other words, and demonstrates to the user how small changes can incrementally increase use of plain language in written health information.

## Methods

### Study design

The study used a two-arm parallel-group randomised-controlled trial study design with participants randomly assigned to intervention or control groups (Figure 1). This trial was prospectively registered with the Australian New Zealand Clinical Trials Registry (ACTRN12623000386639) and approved by the University of Sydney Human Research Ethics Committee (2023/276).

**Figure 1.**
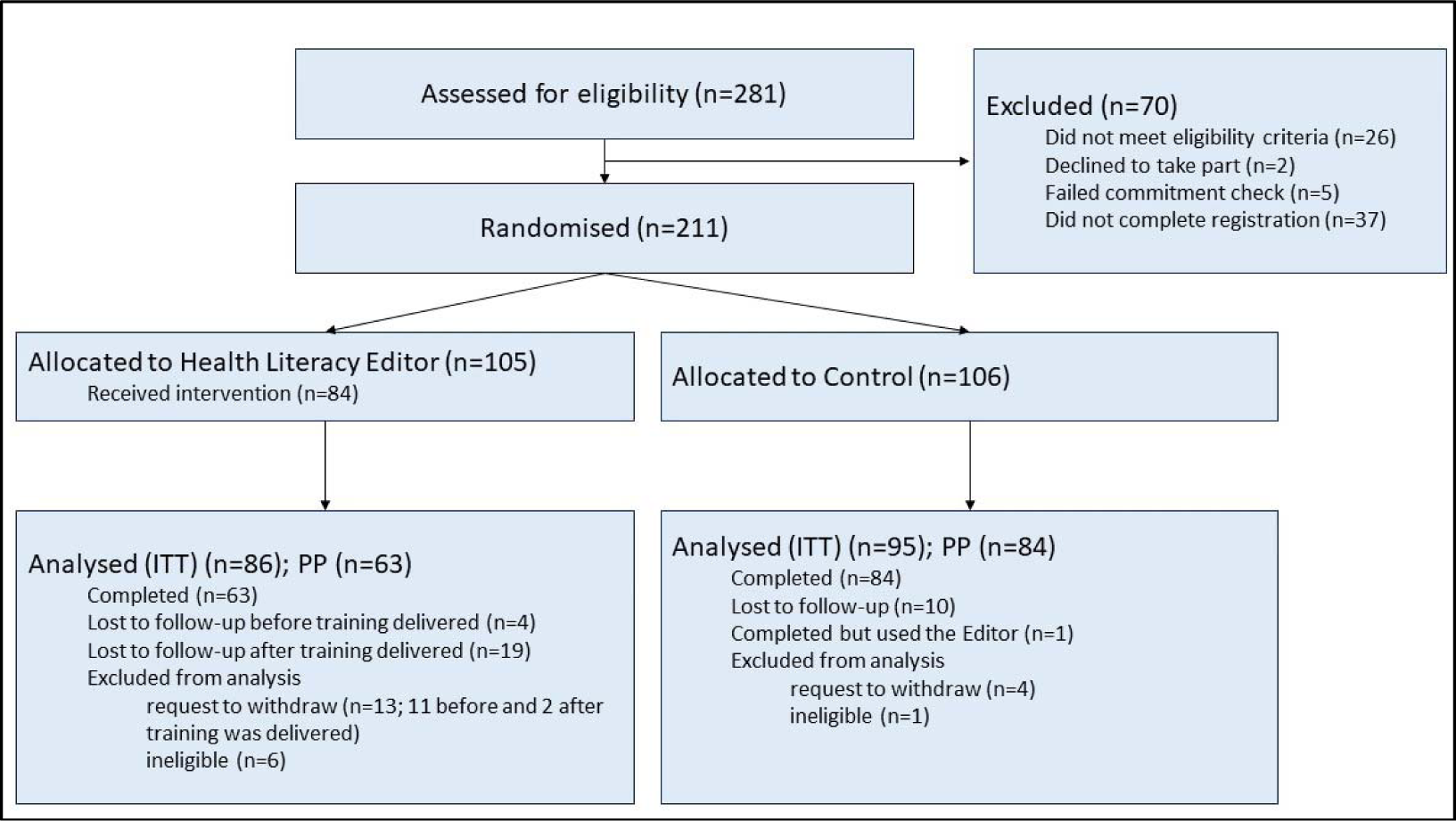
Participant flow and study design.

### Participants

Eligible participants were adults in Australia whose work involved developing health information. Students in medicine, allied health and health sciences were also eligible to take part. Participants also had to positively affirm their commitment to the trial (“Do you commit to providing thoughtful answers in this study?”). Participants were not eligible to take part if they had previous experience using the Health Literacy Editor. Participants were recruited online through health networks, newsletters, and social media.

### Intervention groups

#### Health Literacy Editor group

The Health Literacy Editor is a browser-based software application that gives objective real-time feedback on the complexity of health information. The Health Literacy Editor comprises six assessments: readability as measured by the Simple Measure of Gobbledygook (SMOG) formula, complex language, passive voice, text structure, lexical density and diversity, and person-centred language. These are each presented as global scores, with additional, more specific feedback flagged in the text itself through highlighting individual words and sentences. Full details of the development are published elsewhere (31). User testing with health staff has helped improve the quality of training, instructions, and feedback that the tool provides (32). Participants randomised to this group attended a 30-minute online training session in which they learned how to use the tool. Three training resources were also embedded within the tool: a help page that contained instructions, video tutorials and worked examples; a self-check tool comprising 5 questions to check understanding of key concepts; and a 2-page PDF introducing key concepts. After completing training, participants were instructed to use the Health Literacy Editor to help them revise the three health texts.

#### Control group

Participants were asked to use their usual processes to revise three health texts. No further training was provided.

### Procedure

Participants provided consent online, along with demographics, details about professional or student role, and their experience developing health information. They were then randomised 1:1 to intervention group or control using the Qualtrics survey platform’s Mersenne Twister algorithm. Participants were emailed a link to a second survey which asked them to revise three health texts, each approximately 200 words and written at a Grade 14 reading level, on the topics of dementia, cancer and sciatica (Appendix A). Three topics relating to different commonly occurring health conditions were selected to mitigate bias attributable to topic area expertise. Selection of the texts ensured that there were enough long words and sentences, complex language, and passive voice that participants could demonstrate their ability to simplify the text according to health literacy guidelines.

Instructions asked all participants to revise each text to make it easy for most people to understand, to aim for Grade 8 to 10 reading level, and to retain any key messages within the text. The reading level range was selected to reduce participant burden and ensure the revision task was feasible. To aid revision, a brief description of the purpose for each text was also provided. After revising the texts, participants completed items about self-reported estimates of time taken to revise the text. Participants in the intervention group reported on the features they used, and the tool’s usability and acceptability. Participants received a $50 gift card to thank them for their time.

## Outcomes

### Primary outcome

The primary outcome was grade reading score as measured by SMOG (further detail provided in Box 1)(33).

##### Box 1. Assessment tools incorporated into evaluation outcomes

| Assessment tool | Description |
| --- | --- |
| Grade reading score | Grade reading score provides an estimate of text difficulty that corresponds to the US grade reading level required to understand the meaning of a text. In Australia, health literacy guidelines recommend that information is written at a grade 8 reading level or lower (34).<br>The SMOG formula is widely used in health literacy research (35) and calculates grade reading score based on the number of long words (defined as 3 syllables or more) and the number of sentences (33). The resulting score corresponds to the grade reading level at which you would expect a person to correctly answer 100% of multiple-choice comprehension questions. |
|  | The SMOG score produced by the Health Literacy Editor is closer to the reference standard i.e. hand-calculated scores than SMOG scores produced by other software (36). |
| **SHeLL Health Literacy Editor** | In addition to grade reading score, the Health Literacy Editor produces a score for complex language and passive voice. These assessments were custom-built as there were no other existing validated formulas or programming available.<br>The complex language assessment seeks to estimate the proportion of the text that contains words that may be unfamiliar to readers. The Health Literacy Editor draws from several resources to identify complex language, including a database comprising more than 270 million words from diverse English-language sources to identify words that are uncommon in English (37).<br>The passive voice assessment uses natural language processing to identify patterns of the verb 'to be' (e.g., 'is,' 'were,') and a past participle (e.g. 'delivered', 'given') that indicate passive voice (36). |
| **Patient Education Materials Assessment Tool (PEMAT)** | The PEMAT is a widely used and validated health literacy tool that asks the assessor to subjectively rate whether printed health materials adhere to 24 items (12). This provides an estimate of 'understandability' i.e. how easy a material is to understand; and 'actionability' i.e. how easy a material is to act upon. Topics include content, word choice and style, use of numbers, organisation, layout and design, and use of visual aids. |

### Secondary outcomes

#### Objective text complexity

Using simple everyday language is a key health literacy recommendation to improve the quality of health information (12). This was assessed using the Health Literacy Editor’s ‘complex language score’ (Box 1). The complex language score reports the proportion of words (%) that are uncommon in English, acronyms, or words with a suggested alternative in the tool’s thesaurus (36).

#### Objective passive voice

Using active voice is a key recommendation to improve how easy health information is to understand (12). Passive voice was assessed using the Health Literacy Editor (Box 1).

#### Subjective expert ratings

Two co-investigators with expertise in health literacy (JA and OM) assessed each revised text, masked to intervention group. Scores represent average ratings on a 5-point Likert scale (Strongly disagree [1] to strongly agree [5]). Three ratings for each text were produced. The first two, ‘Content’ and ‘Word choice and style,’ correspond to PEMAT topics (12) (Box 1). The content topic relates to a clear purpose and absence of distracting information. Word choice and style refers to use of common everyday language, minimal and defined medical terms, and use of the active voice where possible. A third rating ‘retained meaning’ reflects whether key messages were retained in the revised texts. This was added to ensure texts were not simplified by removing content.

#### Time to complete text revisions

Participants in both groups were asked to estimate the number of minutes to complete all three revision tasks.

#### Intervention acceptability

Participants in the intervention group provided acceptability ratings via two validated instruments. The System Usability Scale (38, 39) produces a score from 0 (low) to 100 (high). A score of 70 is considered acceptable and a score of 90 or more is considered superior (39). The Technology Acceptance Model (40) comprises two subscales: perceived usefulness and perceived ease of use, with scores ranging from 1 (low) to 5 (high). Scores are predictive of current and future use of a product (40).

#### Intervention engagement

Participants were asked which Health Literacy Editor features they engaged with when revising the text. Participants who reported using at least two of three key features described in training (Readability, Complex language and Passive voice) were assessed as having adequate engagement.

### Analysis

#### Sample size

A sample size estimate of 120 (60 participants per group) was calculated to have 90% power at α = 0.05 to detect a moderate effect size (Cohen’s f = 0.30) in the main outcome (grade reading score). An additional buffer allowed for up to 33% drop-out before the text revision task was completed for a total 180 participants. Sample size was adjusted during recruitment to account for a larger than expected non-completion rate (N=211).

#### Statistical analysis

Univariable regression models analysed differences for resources developed using the Health Literacy Editor (averaged across the three texts) and those developed by participants in the control condition. Intention-to-treat (ITT) and per-protocol (PP) analyses are presented. For ITT, the scores of the original texts were retained as the scores for revised texts for participants who did not complete the revision task. That is, we assumed no changes were made to the text. PP analysis included only participants who submitted revised versions of the texts (for participants in the intervention group this included attendance at training). One participant in the control group who used the Health Literacy Editor was excluded from the PP analysis. Descriptive statistics summarised information about usability, acceptability, and engagement (intervention group only). For all subjective ratings, assessors were unaware of intervention group.

Analyses were performed using IBM SPSS Statistics 26. The threshold for significance of the primary outcome was p<0.05 and all hypothesis tests were 2-sided. The same significance threshold was used for analyses of secondary outcomes and can be interpreted as exploratory.

### Patient and public involvement

A community member was involved in discussions about study design. Methods and outcome measures related to community ratings of the revised texts incorporated their feedback. This subsequent component of the project is not reported in this manuscript and will be reported separately. Several health services staff and university research staff helped pilot and improve the training materials and ensured that the text revision task was feasible without placing undue burden on participants.

## Results

### Sample characteristics

Participants were recruited between May and November 2023, with follow-up completed by February 2024. The number and flow of participants at each stage is shown in Figure 1. Sample characteristics are summarised in Table 1. Participants were on average 41.0 years (SD=11.6), 85.1% identified as female (n=154), 13.3% (n=24) male, 1.7% (n=3) as non-binary or other gender, and most reported working in health services (62.4%, n=113) and government organisations (64.6%, n=117). Characteristics appeared comparable for the PP sample (Table A1).

**Table 1.** Participant characteristics by intervention group, ITT analysis sample (N=181) (n (%) unless otherwise stated)

| Variable | Health Literacy Editor (n=86) | Control (n=95) |
| --- | --- | --- |
| Age; mean (SD) | 41.3 (11.4) | 40.6 (11.8) |
| Gender |  |  |
| Male | 11 (12.8) | 13 (13.7) |
| Female | 74 (86.0) | 80 (84.2) |
| Other gender | 1 (1.2) | 2 (2.1) |
| Role<sup>a</sup> |  |  |
| Student only | 2 (2.3) | 2 (2.1) |
| Staff only | 71 (82.6) | 77 (81.1) |
| Student and staff | 13 (15.1) | 16 (16.8) |
| Engagement (staff only)<sup>a</sup> |  |  |
| Government | 56 (65.1) | 61 (64.2) |
| Health services | 54 (62.8) | 59 (62.1) |
| University or tertiary | 7 (8.1) | 10 (10.5) |
| education |  |  |
| Not-for-profit or charity | 3 (3.5) | 4 (4.2) |
| Consumer advocacy group | 0 (0.0) | 2 (2.1) |
| Industry | 2 (2.3) | 7 (7.4) |
| How often do you develop or revise written health information for patients or the community? |  |  |
| Daily | 16 (18.6) | 19 (20.0) |
| Weekly | 15 (17.4) | 13 (13.7) |
| Monthly | 15 (17.4) | 17 (17.9) |
| A few times a year | 26 (30.2) | 29 (30.5) |
| Once | 2 (2.3) | 1 (1.1) |
| Never | 10 (11.6) | 14 (14.7) |

<sup>a</sup> Multiple roles and engagements could be selected.

Most participants completed follow-up (81.2%, n=147), though this rate was lower for participants in the intervention group compared to control (73.3%, n=63; 89.5%, n=84, respectively).

### Evaluation of revised texts

#### Primary outcome

Compared to text revised by participants in the control group, the texts revised by those in the intervention group had significantly improved grade reading level (Mean Difference (MD)=2.48, 95% CI=1.8413.12, p<0.001, d=0.99; Table 2 and Table A2). Magnitude of effects were larger in PP analysis, with participants in the intervention group reducing the grade reading level by almost 4 grades relative to those in the control group (MD=3.79, 95% CI = 3.2914.28, p<0.001, d=1.58).

**Table 2.**
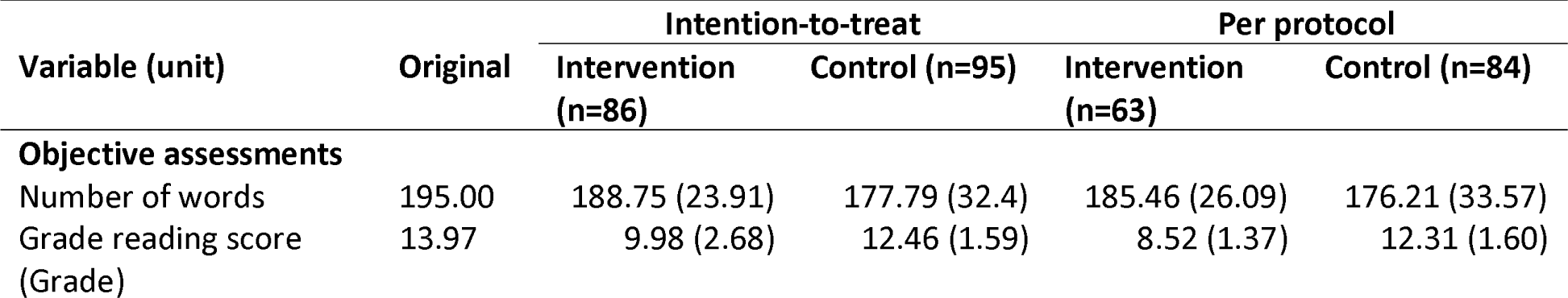
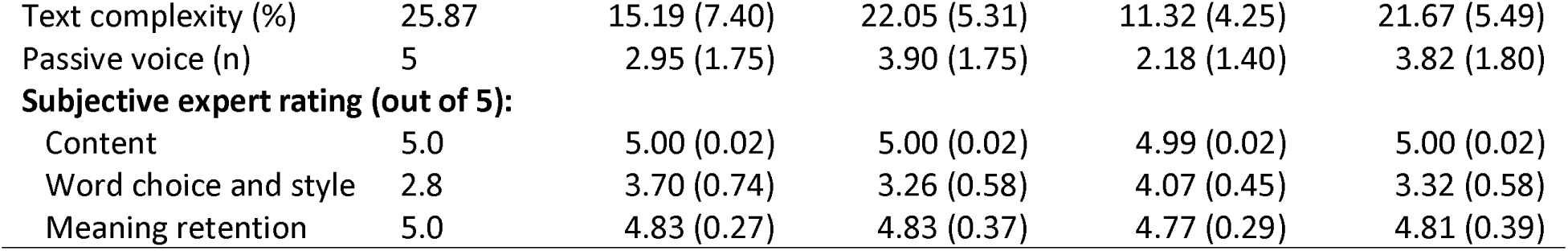
Participant scores for revised materials, by intervention group (N=181) (M (SD))

#### Secondary outcomes

The same pattern was observed for secondary outcomes, with texts revised by those in the intervention group showing lower text complexity (MD=6.86, 95% CI=4.9918.74, p<0.001, d=0.95) and less passive voice (MD=0.95, 95% CI=0.4411.47, p<0.001, d=0.53) (Table 2, Table A2). Expert ratings for word choice and style (common everyday language, minimal and defined medical terms, and active voice) were higher in the intervention compared to control group (MD=0.44, 95% CI = 0.2510.63, p<0.001, d=0.63, Table 2). Magnitude of effects for secondary outcomes were larger in PP analysis, including across both objective and subjective expert ratings. Ratings for content (clear purpose and absence of distracting content) and retaining meaning were high and did not differ significantly across the two groups.

#### Acceptability and engagement

On average, participants rated the Health Literacy Editor an acceptable product that was useful and easy to use (Table 3). Participants using the Health Literacy Editor reported spending an average 65.40 minutes revising the three texts (SD=33.02), compared to an estimated 30.13 minutes for the control group (SD=18.28). Almost all participants (n=59, 93.7%) reported using all three of the Health Literacy Editor’s key assessments: readability, complex language and passive voice.

**Table 3.** Acceptability and engagement with the Health Literacy Editor, n=63.

| Characteristic | Mean (SD) |
| --- | --- |
| System Usability Scale (0 low to 100 high) | 70.99 (13.69) |
| Technology acceptance model (1 low to 5 high) |  |
| Perceived usefulness | 3.76 (0.71) |
| Perceived ease of use | 4.03 (0.59) |
| Time to complete revisions (mins) | 65.40 (33.02) |
|  | **N (%)** |
| Self-reported Health Literacy Editor features used to revise the text |  |
| Readability | 62 (98.4) |
| Complex language | 63 (100.0) |
| Passive voice | 59 (93.7) |
| All three features | 59 (93.7) |

## Discussion

Health information that was revised using the Health Literacy Editor more closely aligned with health literacy and plain language guidelines compared to texts revised according to participants’ standard processes. These texts had a lower grade reading score and used less complex language and passive voice, showing greater potential to meet the health literacy needs of the population, including people who are older, who have had less opportunity for education, and who speak English as a second language. Subjective ratings from health literacy experts provided further evidence that these revised texts were clear and retained the original meaning. Though participants who used the Health Literacy Editor took longer to revise the texts, we believe this time investment is reasonable given the magnitude of effects, the likelihood that participants may become faster with repeated use of the tool, and its strong capability to support scalable and easily accessible health literacy training.

These findings highlight that innovative new tools can meaningfully contribute to bridging the well-documented gap between health literacy policy and practice (1, 2, 6–9). To date, several promising tools have been developed (24–30), with very limited evaluation of their effectiveness (29, 30). This is the first study to use a randomised-controlled trial study design, to show that health literacy software providing objective, real-time and fine-grained feedback on words and sentences is effective in supporting health information providers to develop plain language written materials. Coupled with sound user acceptability ratings, further work is now needed to explore how tools such as Health Literacy Editor can be implemented at scale within an organisation and to evaluate its impact on patient outcomes.

Online tools are well placed to support consistent and scaled uptake of health literacy guidelines by workforce who may have received little training in the area. This is important given that developing health information can be an intermittent activity for clinicians and health staff, particularly when roles are transient or project based. Online tools and training have the advantage of being easily accessed online without geographic or time constraints.

Developing health texts will always need human oversight and expertise. We envisage that health information providers would use the Health Literacy Editor in combination with other tools and strategies whilst maintaining existing quality and safety processes for clinical oversight. These caveats are likely to continue to apply, even with advances in artificial intelligence that may allow these tools to quickly and coherently simplify health information (41, 42).

In addition to the randomised-controlled trial design, several other aspects further strengthen the study findings. For example, though readability is an appropriate primary outcome measure, it has been criticised as a narrow indicator of plain language (43). This study addressed this issue by including a variety of objective and subjective assessments of plain language (complex language and passive voice) and was therefore able to show consistent patterns across a wide range of outcomes. Study findings were also strengthened by asking participants to submit three revised texts on different health topics. This reduced the likelihood that content-area expertise would influence results.

There are also some limitations to this study. Several participants completed intervention training but did not submit revised texts. It is possible that some were overwhelmed by the training or did not see value in the tool if they were already confident in their skills. Qualitative and co-design research may further improve the training and help set appropriate expectations for using the tool. It is also unclear whether results generalise to health information developers in non-government sectors, given the low number of participants from industry, consumer advocacy groups, and tertiary institutions. Secondly, it is unclear whether improved uptake of plain language will translate to improved perceptions of the health information by consumers and patients. Further work is underway to explore whether consumers prefer and can more easily understand texts developed using the Health Literacy Editor. This work may also help understand the relative importance of each objective assessment.

The findings from this study demonstrate that the Health Literacy Editor can support users to apply health literacy and plain language strategies to written text, while retaining key content and meaning. New technologies may make an important practical contribution to achieving the goals set out by health literacy policy for clear health communication, improved health equity, and better health outcomes. These tools have potential to improve health outcomes for people with lower health literacy.

## Data Availability

Deidentified data will be made available on reasonable request.

## Acknowledgements

We would like to acknowledge the support and contributions of Nancy Zaki, Allison Sigmund, Susan Dyer, Dianna Smith-McCue, and Debra Letica in this study. Thank you also to Olivia Mac, Melody Taba, Katie McFadden, and Jenna Smith for their support in piloting the text revision tasks.

## Transparency statement

Julie Ayre affirms the manuscript is an honest, accurate and transparent account of the study being reported. No important aspects of the study have been omitted. Any discrepancies from the study as originally planned have been explained.

## Data sharing statement

Deidentified data will be made available on reasonable request.

