## Appendix A for "Effects of an online plain language tool on health information quality: A randomised controlled trial"

### Appendix A: Original texts

#### Text 1: Sciatica

This text is from a website that gives readers information about **sciatica**, including information about when they should seek immediate medical attention, and information suggesting that they may not need imaging for sciatica.

Sciatica is a condition that can lead to pain in the back and legs. Sciatica is felt as nerve pain radiating from the buttock down the back of the leg, often when sitting, sneezing, coughing or going to the toilet. You may also feel lower back pain, and/or tingling, pins and needles, numbness or weakness in your leg.

Although sciatica pain can be severe, most people find their symptoms improve in time. As sciatica is seen as a more serious low back condition, it's advisable to see your doctor within the first few days of getting the symptoms. To diagnose sciatica, the doctor will take a medical history and examine your spine and legs.

You should seek medical attention immediately if you have problems controlling your bladder or bowels, or have weakness, numbness or severe pain.

The Royal Australian and New Zealand College of Radiologists recommends that an x-ray or other imaging in response to low back pain is only needed if you have other significant symptoms. If you have sciatica, discuss with your doctor whether imaging is required, which it may or may not be, depending on the circumstances.

#### Text 2: Dementia

This text is from a website that gives readers information about **dementia**, including a description of its burden, the symptoms, and risk factors.

Dementia is currently the seventh leading cause of death among all diseases and one of the major causes of disability among older people worldwide.

Dementia is a syndrome that is usually of a chronic or progressive nature. It leads to deterioration in cognitive function beyond what might be expected from the usual consequences of biological ageing. It affects memory, thinking, orientation, comprehension, calculation, learning capacity, language, and judgement. The impairment in cognitive function is commonly accompanied, and occasionally preceded, by changes in mood, emotional control, behaviour, or motivation.

Although age is the strongest known risk factor for dementia, it is not an inevitable consequence of biological ageing. Further, dementia does not exclusively affect older people. Young onset dementia (defined as the onset of symptoms before the age of 65 years) accounts for up to 9% of cases. Studies show that people can reduce their risk of dementia by being physically active, not smoking, avoiding harmful use of alcohol, controlling their weight, eating a healthy diet, and maintaining healthy blood pressure, cholesterol and blood sugar levels. Additional risk factors include depression, social isolation, low educational attainment, cognitive inactivity and air pollution.

#### Text 3: Cancer stages

This text is from a website that gives readers information about **cancer stages**, and why this concept is important for people with cancer.

What does the 'stage' of a cancer mean?

Once a cancer is diagnosed, it needs to be carefully measured in both its size and extent of spread. This is classified through staging. The main factors affecting the stage of a cancer are the size of the primary tumor (where the cancer started), the involvement of lymph nodes, and the potential spread to other organs in the body. Staging is commonly done through a physical examination, imaging studies, and sometimes additional biopsies.

Why do we stage a cancer?

Staging is performed for several reasons. First, prognosis, which is the likely clinical outcome, is highly associated with the stage of a cancer and gives an opportunity to understand life expectancy and, in some cases, the chance for cure.

Second, certain therapies are indicated for some stages but not others. For example, surgery is routinely recommended for earlier stages of cancer, when the cancer has not spread or has only spread to nearby lymph nodes. In contrast, advanced cancer that has spread to distant parts of the body is commonly treated with medications that treat disease throughout the entire body.

Finally, clinical trials that test new drugs are available to patient populations with a specific cancer and stage.
