## Appendix B for "Effects of an online plain language tool on health information quality: A randomised controlled trial"

**Table A1. Participant characteristics by intervention group, PP analysis sample (N=147) (n (%) unless otherwise stated)**

| **Variable** | **Health Literacy Editor (n=63)** | **Control (n=84)** |
| --- | --- | --- |
| Age; mean (SD) | 40.7 (11.0) | 40.0 (11.3) |
| Gender |  |  |
| Male | 8 (12.7) | 11 (13.1) |
| Female | 54 (85.7) | 71 (84.5) |
| Other gender | 1 (1.6) | 2 (2.4) |
| Role^a^ |  |  |
| Student | 11 (17.5) | 16 (19.0) |
| Staff | 61 (96.8) | 82 (97.6) |
| Government | 41 (65.1) | 54 (64.3) |
| Health services | 38 (60.3) | 51 (60.7) |
| University of tertiary education | 7 (11.1) | 9 (10.7) |
| Not-for-profit or charity | 3 (4.8) | 3 (3.6) |
| Consumer advocacy group | 0 (0.0) | 1 (1.2) |
| Industry | 1 (1.6) | 6 (7.1) |
| How often do you develop or revise written health information for patients or the community? | | |
| Daily | 8 (12.7) | 17 (20.2) |
| Weekly | 14 (22.2) | 11 (13.4) |
| Monthly | 11 (17.5) | 15 (18.3) |
| A few times a year | 19 (30.2) | 27 (32.9) |
| Once | 1 (1.6) | 1 (1.2) |
| Never | 8 (12.7) | 11 (13.1) |

^a^Multiple roles could be selected.

**Table A2. Effect size estimates**

|  | **Intention-to-treat** | | | **Per protocol** | | |
| --- | --- | --- | --- | --- | --- | --- |
| **Variable (unit)** | **Mean difference (95% CI)** | **Cohen’s d** | **P value** | **Mean difference  (95% CI)** | **Cohen’s d** | **P value** |
| Grade reading score (Grade) | 2.48 (1.84 to 3.12) | 0.99 | <0.001 | 3.79 (3.29 to 4.28) | 1.58 | <0.001 |
| Text complexity (%) | 6.86 (4.99 to 8.74) | 0.95 | <0.001 | 10.35 (8.71 to 12.00) | 1.45 | <0.001 |
| Passive voice (n) | 0.95 (0.44 to 1.47) | 0.53 | <0.001 | 1.64 (1.10 to 2.18) | 0.90 | <0.001 |
| Subjective expert rating |  |  |  |  |  |  |
| Content | 0.00 (-0.01 to 0.01) | -0.02 | 0.902 | -0.00 (-0.01 to 0.06) | 0.00 | 0.720 |
| Word choice and style | 0.44 (0.25 to 0.63) | 0.63 | <0.001 | 0.75 (0.58 to 0.93) | 1.16 | <0.001 |
| Meaning retention | 0.00 (-0.09 to 0.10) | 0.00 | 0.977 | -0.04 (-0.16 to 0.07) | -1.83 | 0.493 |
